# Identifying the risk profile of anemia subtypes and hemodynamic obstetric complications in relation to peripartum cardiomyopathy

**DOI:** 10.64898/2026.06.11.26355493

**Authors:** Kayla Sompel, Elise Shalowitz, Melinda Davis, Elizabeth Kudron, Shannon Son, David Kao

## Abstract

**Background:** Peripartum cardiomyopathy (PPCM) is a leading cause of maternal mortality worldwide, with worse outcomes associated with African Ancestry and delayed presentation. However, the mechanisms underlying PPCM are incompletely understood.

**Objective:** Use a large, nationwide cohort to explore associations between PPCM and underexplored perinatal risk factors and complications of childbirth.

**Methods:** Public hospital discharge data were obtained from eleven U.S. states between 2003-2019. Delivery hospitalizations, patient characteristics and obstetric complications were identified using ICD-9 and -10 CM codes. Only cases with unique patient identifiers enabling readmission analysis were included. The primary outcome was incident PPCM coded between 30 days antepartum and 150 days postpartum.

**Results:** Of 7,424,916 delivering patients, 5,488 patients were diagnosed with PPCM. Patients with PPCM had higher rates of anemia, anemia of chronic disease (ACD), iron deficiency anemia (IDA), sickle cell disease (SCD), sickle cell trait (SCT), red blood cell (RBC) transfusion, and postpartum hemorrhage (PPH) (p<0.001 for all). Transfusion was associated with increased risk of postpartum PPCM both with PPH (OR 2.16) and without PPH (OR 1.92). In multivariable analysis antepartum diagnosis was more common in the setting of anemia (OR 2.5, p<0.001), ACD (OR 16.31 p<0.006), SCD (9.11, p=0.002) and SCT (OR 2.96 p=0.021), while IDA was associated with peripartum and postpartum PPCM (OR 2.04 p<0.001)

**Conclusion:** Anemia subtypes, RBC transfusion, and PPH were associated with an increased risk for PPCM. The magnitude of risk varied by race and timing of presentation. Further study of peripartum interventions directed at these risk factors is warranted.

## Introduction

Peripartum cardiomyopathy (PPCM) is a life-threatening complication of pregnancy that remains a leading cause of maternal mortality globally, particularly in low-middle income countries (1–3). PPCM is defined as nonischemic, systolic heart failure, with a left ventricular ejection fraction (LVEF) of less than 45%, that occurs in individuals towards the end of pregnancy or the early months following delivery that cannot be explained by an alternative etiology (4, 5),(6). The incidence of PPCM has geographic(7) and racial variation(8–10) with rates as high as 1 in 102 births in Nigeria (11, 12) to as low as 1 in 20,000 births in Japan(13). In the United States (US), PPCM disproportionately affects people of African Ancestry (AA), with PPCM rates as high as 1 in 664 for AA patients compared with 1 in 2450 non-Hispanic white patients and 1 in 6729 Hispanic patients (7). The clinical course of PPCM is highly variable with clinical presentations ranging from mild symptoms and a short interval to complete LVEF recovery, to rapidly progressive disease and devastating outcomes. The high morbidity and mortality rates of PPCM have remained largely unchanged over the past 50 years (1) (7, 9, 14–16) and PPCM continues to account for 5% of heart transplants in females in the United States(17).

In addition to AA race, previously identified risk factors for PPCM include chronic hypertension (cHTN), hypertensive disorders of pregnancy (HDP), advanced maternal age (AMA), diabetes mellitus (DM), gestational diabetes mellitus (GDM), substance use disorder (SUD), obesity, multiparity, cesarean delivery (CD), mood disorder, asthma, autoimmune disease (18, 19) genetic predisposition(20, 21) high social vulnerability index (SVI)(8, 22), and anemia(4, 19). Unrelated to pregnancy, anemia is a known risk factor and comorbidity associated with heart failure. Various anemia subtypes including sickle-cell disease (SCD)(23) and trait (SCT) (24), iron deficiency anemia (IDA)(25), red blood cell (RBC) transfusions(26, 27), hemorrhage(28, 29), and thalassemia(30) to increase heart failure morbidity and mortality. While these risk factors have been associated with higher risk of heart failure, they have not been systematically characterized in relation to PPCM risk. Furthermore, the impact of complications of childbirth related to hemodynamic instability such as postpartum hemorrhage (PPH), red blood cell (RBC) transfusion, and unscheduled Cesarean section (CS) has not been thoroughly investigated.

Despite the identification of risk factors for PPCM and efforts to create a PPCM clinical risk stratification score(19), there are currently no definitive clinical parameters to prompt diagnostic evaluation of PPCM during pregnancy, nor are there efficient strategies for identifying high-risk patients to follow postpartum(19). PPCM symptoms often overlap with normal physiologic changes during and after pregnancy including dyspnea on exertion, lower extremity edema, and fatigue which can lead to missed or delayed diagnoses and ultimately worsen prognosis (5). Additional data is clearly needed to more adequately develop clinical protocols to prompt early diagnosis, intervention, and possibly even prevention to improve PPCM outcomes. Because the impact of PPCM is greater in LMIC, it is particularly important that risk models be implementable in a wide range of clinical settings.

We utilized a large, 17-year database (2003-2019) of hospitalizations from across the United States to identify additional risk factors and describe the race-based differences in the predictors of PPCM among delivering patients. We hypothesized that risk factors that impact hemodynamic stability such as PPH, RBC transfusion, hemoglobinopathies, IDA, and unscheduled CD would be associated with a higher risk of PPCM and that the impacts would vary significantly between racial groups. While clinical informatics is limited by the inability to review key diagnostic criteria and medical management for individual cases, this study aims to utilize aggregate data to identify clinically relevant associations in a rare disease to serve as a framework for future research studies and optimization of clinical decision-making tools geared toward early diagnosis and intervention of PPCM.

## Methods

### Data sources

Deidentified hospital records were obtained from either the Agency for Healthcare Research and Quality Healthcare Utilization Project Statewide Inpatient Database (Arkansas (2015-9), Arizona (2003-19), Colorado (2015-19), Florida (2004-19), Louisiana (2010-2019), Maryland (2016-9), North Carolina (2015-9), New York (2015-9), South Dakota (2015-9), Washington 2003-19), and Wisconsin (2015-2019). In total, there were 20,502,320 hospital records. Records that reported ages < 15 years old or > 50 years old or male sex were excluded. Childbirth was identified based on ICD-9/10 diagnosis and procedures codes (see Supplemental material). Ethical oversight was provided by COMIRB exemption from the University of Colorado (#08-1258) on February 26, 2009.

Patients were classified as having PPCM using ICD-9 CM (674.50-54) and ICD-10 CM codes (O903), non-PPCM patients were classified by absence of these codes. Timing of PPCM presentation was defined based on timing of hospitalization relative to delivery using unique patient identifiers generated by the distributors of data. Specifically, a hospitalization was classified as antepartum if PPCM diagnosis was made between ≥30 days prior to hospitalization for delivery. Peripartum presentation was defined as diagnosis during delivery hospitalization. Postpartum presentation was defined as rehospitalization for PPCM within 5 months (≤ 150 days) following a delivery where PPCM was not noted at the time of delivery. Timing of hospitalization to identify readmissions were based on date stamps assigned by the data sources. The window of the last month of pregnancy up to 5 months postpartum was selected based on the definition described by NHLBI(31).

Demographics and outcomes for each hospitalization were determined using dataset documentation. Advanced maternal age (AMA) was defined as ≥ 30 years old, as some datasets only provided age data in 10-year intervals. Race and ethnicity were stratified as non-Hispanic white, AA, Hispanic, Asian, Native American (NA) and Other (all remaining). Comorbid conditions and procedures performed were identified using ICD-9 and ICD-10 CM codes. Hypertensive disorders of pregnancy (HDP) were a composite definition of a history of gestation hypertension (642.3), preeclampsia (642.4, 642.5), or eclampsia (642.6). Chronic HTN complicating pregnancy was considered a separate comorbid condition (642.0, 642.1, 642.2). Remaining ICD-9 and ICD-10 CM codes used for each diagnosis or procedure for the present analysis are available in the online supplement.

#### Statistical Analysis

Rates of PPCM per delivery overall and by race were estimated in terms of PPCM cases/1000 births. Analyses were performed using a chi-square test for categorical variables and Student’s t-test for continuous variables. Associations between PPCM and potential risk factors were determined using logistic regression. The multivariable analysis predicting PPCM included covariates present at the time of delivery: maternal age ≥ 30 years, race, ethnicities, gestational diabetes mellitus, HDP, preeclampsia/eclampsia, premature labor, cesarean section, PPH, RBC transfusion, SCT, sickle cell disease, SCD, alpha and beta thalassemia, chorioamnionitis, and influenza. We applied our previously-derived PPCM risk score to the present data as a validation study(19). We then assessed incremental value of complication of delivery using multivariable analysis and analysis of variance (ANOVA) for interaction terms. Statistical significance was set at a p-value of <0.05.

Data were managed and harmonized using Google Cloud Storage and Google BigQuery. All statistical analyses were performed using Stata (StataCorp, College Station, TX) version 14.1, RStudio (version 2025.09.2+418, Posit Software, Boston, MA) and R Statistical Package (R Project for Statistical Computing, Vienna, Austria) version 4.4.1. R packages corresponding to different aspects of analysis as detailed in the on-line supplement. Code is available from the authors on reasonable request.

## Results

In total 20,241 PPCM records and 20,482,079 hospital-based deliveries were identified corresponding to 1 PPCM hospitalization per 1013 deliveries (**Table 1**). Where race was reported, the majority of delivering patients were non-Hispanic white (55.7%) followed by AA (12.0%), Hispanic (10.7%), Asian (5.0%), and NA (0.8%). The PPCM cohort was primarily white (41.7%), but a larger percentage was non-white; 38.6% AA, 7.2% Hispanic, 2.7% Asian, and 1.3% NA (, p < 0.001). Rates of PPCM varied by race, with AA and NA patients having the highest prevalence of PPCM with 1 in 308 births and 1 in 641 births respectively, compared to 1 in 1,321 births in white patients, 1 in 1,466 births in Hispanic patients, and 1 in 1,821 in Asian patients (p< 0.001). Patients with PPCM were more frequently over 30 years of age (59.5% vs. 43.2%, p < 0.001) and had higher rates of comorbidities including DM, cHTN, anemia, asthma, tobacco use, and SUD (p < 0.001 for all). Using our previously validated PPCM risk score, (19) we found that peope with PPCM had a significantly higher aggregated risk compared to their non-PPCM counterparts (24.6% vs. 3.0% with risk score > 3).

**Table 1:** Deliveries with no PPCM vs. all PPCM hospitalizations with and without delivery, all regions, N (%)

| Characteristic | Deliveries<br>(N=20,482,079) | PPCM<br>Yes (N=20,241) | p |
| --- | --- | --- | --- |
| Maternal age > 30 years | 8845323 (43.2) | 12035 (59.5) | <0.001 |
| **Race** |  |  |  |
| White | 10723998 (55.7) | 8127 (41.7) | <0.001 |
| AA | 2316275 (12.0) | 7522 (38.6) |  |
| Hispanic | 2056191 (10.7) | 1405 (7.2) |  |
| Asian | 958183 (5.0) | 527 (2.7) |  |
| Native American | 159240 (0.8) | 258 (1.3) |  |
| Other | 3026516 (15.7) | 1653 (8.5) |  |
| **Comorbidities** |  |  |  |
| Diabetes mellitus | 220362 (1.1) | 1813 (9.0) | <0.001 |
| Chronic hypertension | 1510979 (7.4) | 9433 (46.6) | <0.001 |
| Anemia | 2312201 (11.3) | 6969 (34.4) | <0.001 |
| Asthma | 619657 (3.0) | 2063 (10.2) | <0.001 |
| Smoker | 771929 (3.8) | 3002 (14.8) | <0.001 |
| Substance abuse | 472259 (2.3) | 2025 (10.0) | <0.001 |
| PPCM risk score > 3 | 611913 (3.2) | 4972 (25.5) | <0.001 |

### PPCM cases including rehospitalizations

There were a total of 6,665,871 records reporting outcome of delivery that had state-assigned unique patient identifiers allowing linkage to hospitalization prior to or following childbirth. Of these deliveries, 5493 were found to have hospitalizations reporting PPCM that occurred between one month before antepartum to 5 months after hospitalization for delivery. (∼1/1213 deliveries). Over 80% of cases occurred within 14 days following delivery **(Figure 1).** Antepartum presentation (hospitalization with PPCM prior to delivery hospitalization) comprised 100 (1.8%) of PPCM cases, whereas 3756 (68.3%) patients were hospitalized with PPCM after a normal postpartum discharge. Characteristics of people with no PPCM are compared to patients with PPCM presenting ante- vs. peri- vs. postpartum in **Table 2a**. Distributions of PPCM presentation according to race are summarized in **Table 3**. AA patients were most likely to present with PPCM post-partum (73%) whereas NA patients were least likely (50.0%, p< 0.001). Antepartum-presenting Patients with PPCM had higher rates of chronic comorbidities including DM, cHTN, anemia, asthma, tobacco use, and SUD when compared to their PP counterparts (p< 0.001 for all). Interestingly, ante-and peripartum presenting Patients with PPCM were more likely to have elevated PPCM risk scores compared to their postpartum-presenting counterparts (26.8% vs. 31.5% vs. 17.0%, respectively with risk score > 3, p<0.001). Furthermore, ante- and peripartum-presenting Patients with PPCM had higher rates of any hypertensive disorders in comparison to postpartum-presenting patients with PPCM (48.0% vs. 54.7% vs. 37.8%, p<0.001), although all were higher than patients without PPCM (11.4%). This included cHTN (35.% vs. 34.3% vs. 20.2%) and preeclampsia/eclampsia (23.0% vs. 36.0% vs. 20.3%). However, gHTN was more common in postpartum-presenting patients (9.5%) compared with ante- (6.0%) or peripartum-presenting patients (9.0%) (p<0.001 for all).

**Figure 1.**
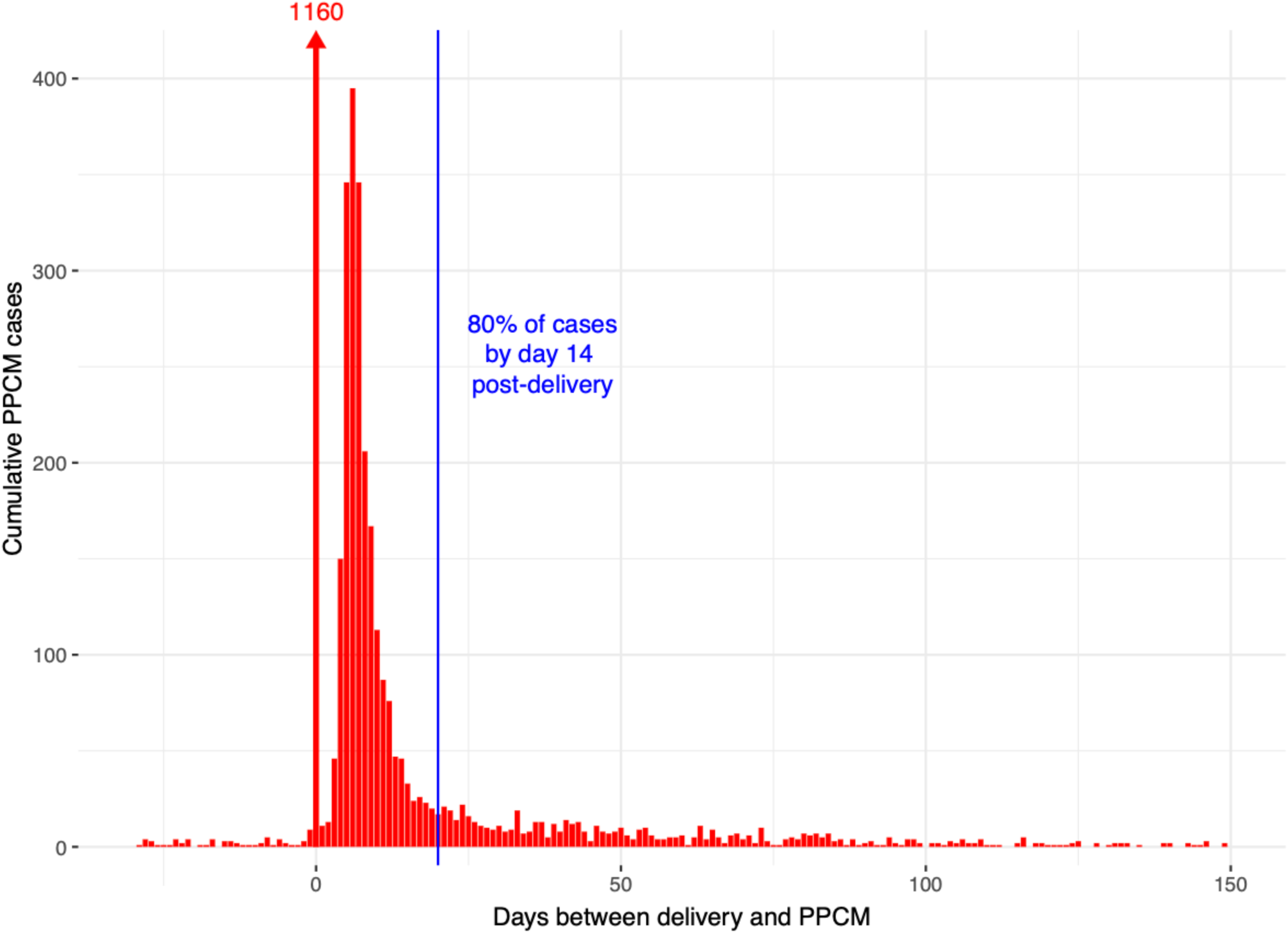
Cumulative PPCM cases according to time of presentation in relation to delivery. Cumulative incidence of peripartum cardiomyopathy (PPCM) cases plotted as a function of time relative to delivery. Cases are stratified by timing of presentation: antepartum (before delivery), peripartum (at the time of delivery), and postpartum (following delivery). The figure illustrates the distribution of PPCM diagnoses across the peripartum continuum, highlighting that the majority of cases present in the postpartum period.

**Table 2:**
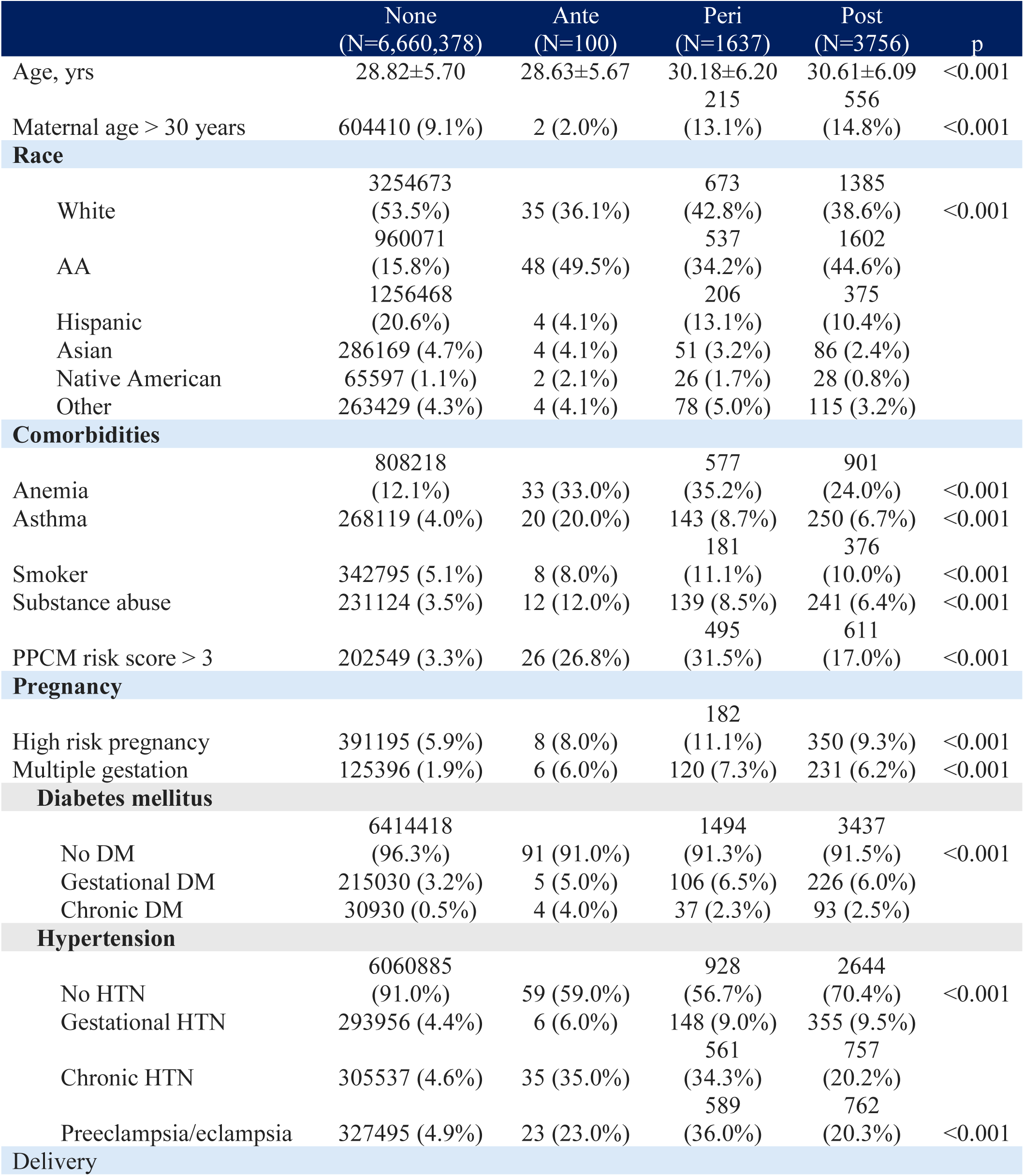
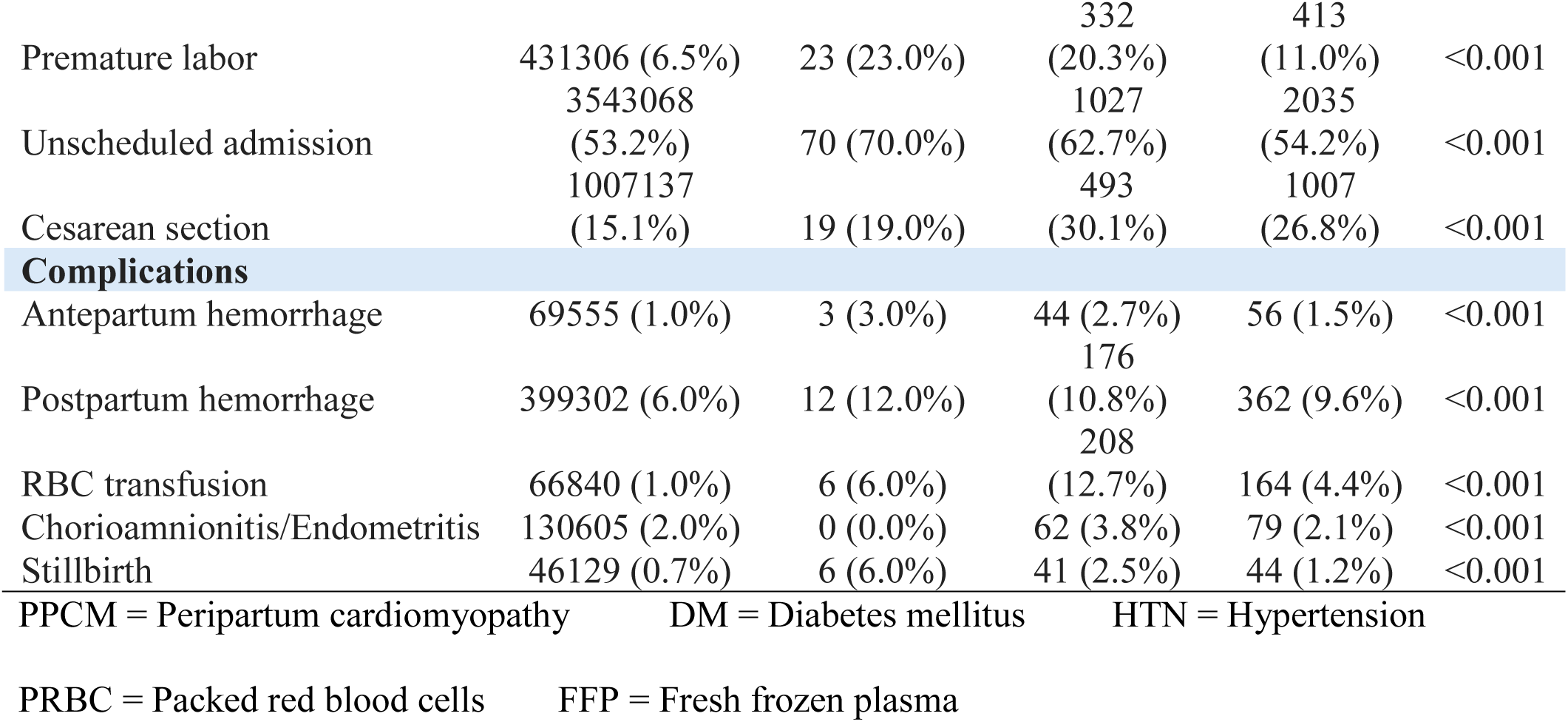
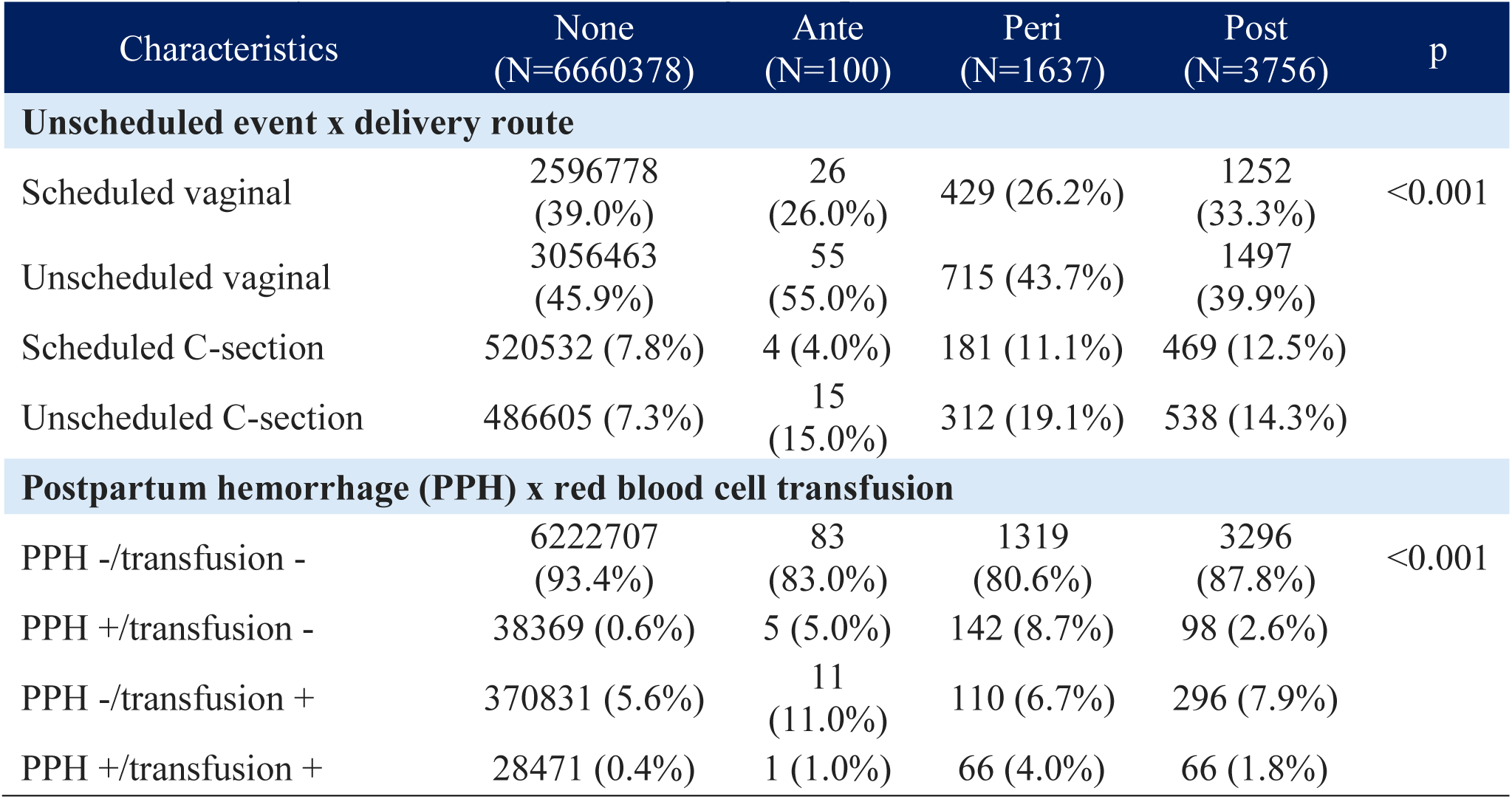
Characteristics of delivering patients with unique patient identifiers by timing of PPCM presentation. Table 2b Delivery conditions and hemorrhagic complications

**Table 3.** Timing of PPCM presentation by race.

| PPCM timing | White<br>(N=2093) | AA<br>(N=2187) | Hispanic<br>(N=585) | Asian<br>(N=141) | Native<br>Am.<br>(N=56) | Other<br>(N=197) |
| --- | --- | --- | --- | --- | --- | --- |
| Antepartum | 35 (1.7%) | 48 (2.2%) | 4 (0.7%) | 4 (2.8%) | 2 (3.6%) | 4 (2.0%) |
| Peripartum | 673 (32.2%) | 537 (24.6%) | 206<br>(35.2%) | 51<br>(36.2%) | 26<br>(46.4%) | 78 (39.6%) |
| Postpartum | 1385<br>(66.2%) | 1602<br>(73.3%) | 375<br>(64.1%) | 86<br>(61.0%) | 28<br>(50.0%) | 115<br>(58.4%) |

### Delivery complications

Delivery characteristics and complications of antepartum and postpartum presenting Patients with PPCM are summarized in **Table 2b**. Premature labor, unscheduled delivery, CD, hemorrhagic complications, blood transfusion, and inflammatory uterine conditions were all more common in patients who developed PPCM than those without. Higher rates of premature labor and unscheduled hospitalization were observed in ante- and peripartum presentations than postpartum. Distribution of delivery conditions (scheduled/unscheduled vs. vaginal/Caesarean delivery) are summarized in **Figure 2**. CD was more commonly observed in patients developing PPCM than those without, with unscheduled CD having the greatest difference compared to people without PPCM. These differences appeared greatest in patients presenting peripartum. While we could not discern whether PPCM was discovered before or after delivery in peripartum cases, but CD was more frequently associated with postpartum than antepartum presentation. However, given the high rates of PPCM presenting at a subsequent rehospitalization following delivery (82.2%), operative delivery appears to be highly associated with postpartum PPCM accounting for every 1 in 1111 scheduled CDs and 1 in 917 unscheduled CDs (**Figure 3**).

**Figure 2.**
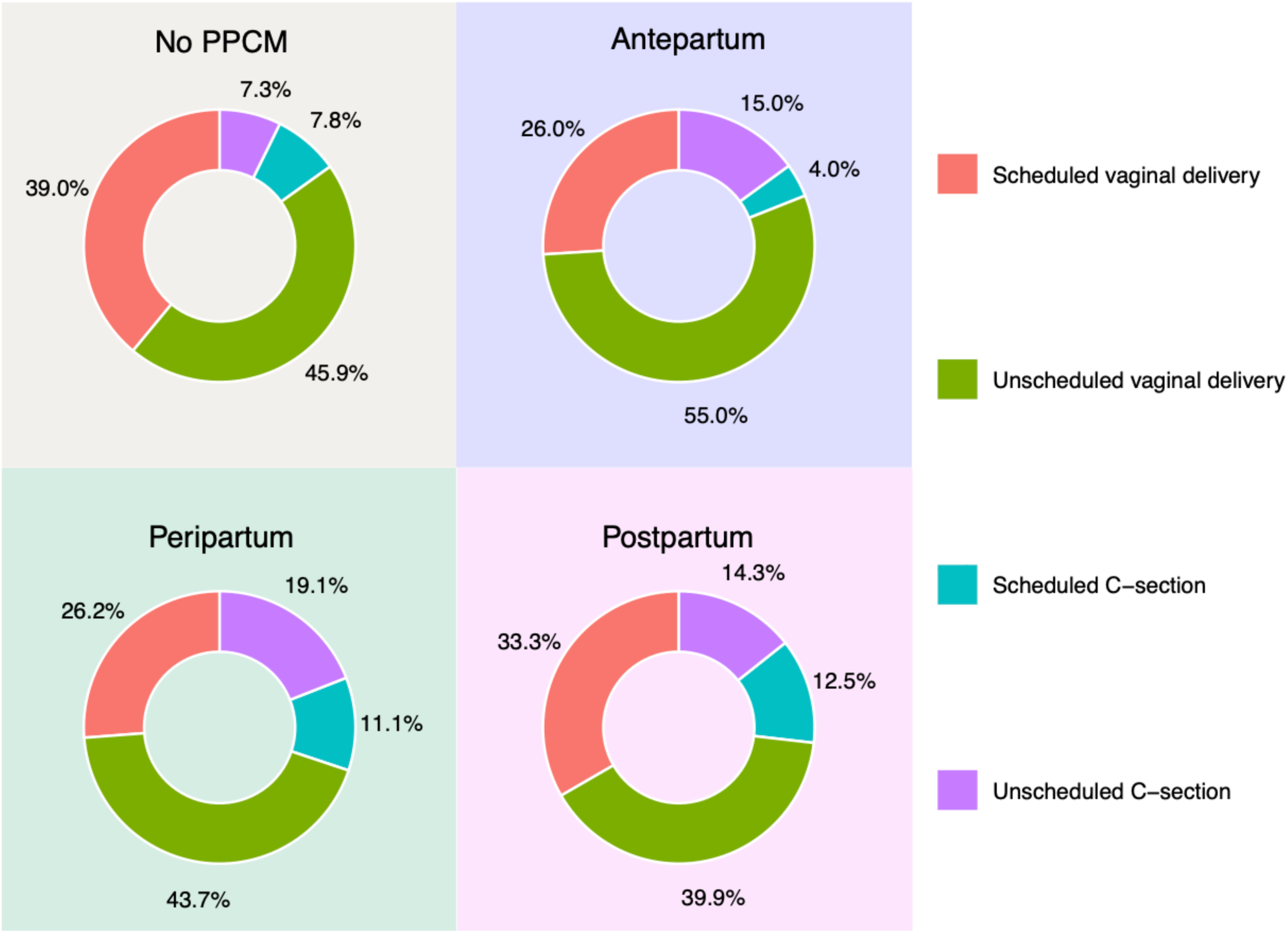
Distribution of scheduled vs. unscheduled vaginal vs. caesarean section delivery according to PPCM presentation. Bar graph depicting the proportion of delivery modes — scheduled vaginal, unscheduled vaginal, scheduled cesarean section, and unscheduled cesarean section — stratified by PPCM presentation timing (no PPCM, antepartum, peripartum, and postpartum). Compared to patients without PPCM, those with peripartum and postpartum PPCM demonstrated higher rates of cesarean delivery and unscheduled admissions. All comparisons were statistically significant (p < 0.001).

Rates of antepartum and postpartum hemorrhage were higher in all presentations of PPCM compared with patients without PPCM. Rates of hemorrhage were highest in antepartum-and lowest in postpartum-presenting patients with PPCM. Transfusion was notably more common in patients presenting peripartum. On further stratification (**Table 2b**), both PPH alone and transfusion alone were more common in patients with PPCM than those without. Our antepartum PPCM cohort also had higher rates of uterine infection (3.6% vs. 2.1%) and stillbirth (2.8% vs. 1.1%) compared to non-Patients with PPCM (p<0.001 for both).

### Multivariable risk factor analysis

Multivariable odds of factors associated with PPCM are summarized in **Figure 3a** (pre-delivery characteristics) and **Figure 3b** (delivery conditions and complications). Numeric odds ratios are available in the on-line supplement (**Table S1**). Due to our focus on potential contributors to PPCM development, multivariable analysis was performed with pregnancy characteristics for all presentations, whereas delivery conditions and complications were only analyzed for peri- and postpartum presentation. There were several differences in magnitude of multivariable risk between ante-, peri-, and postpartum PPCM presentation for multiple risk factors. Odds were highest for antepartum followed by peripartum and lowest in postpartum PPCM for anemia (OR 2.54 [1.65-3.90], OR 2.16 [1.93-2.43], OR 1.43 [1.32-1.55] respectively, p<0.001 for all and chronic hypertension (OR 8.51 [5.47-13.2], OR 4.24 [3.67-4.91], OR 2.54 [2.29-2.83] respectively, p<0.001 for all). Odds associated with age > 30 years were highest in postpartum- (OR 1.56 [1.45-1.66], p<0.001) followed by peripartum- (OR 1.27 [1.15-1.41], p<0.001) and nonsignificant for antepartum-presenting patients with PPCM (p=0.693).

**Figure 3a.**
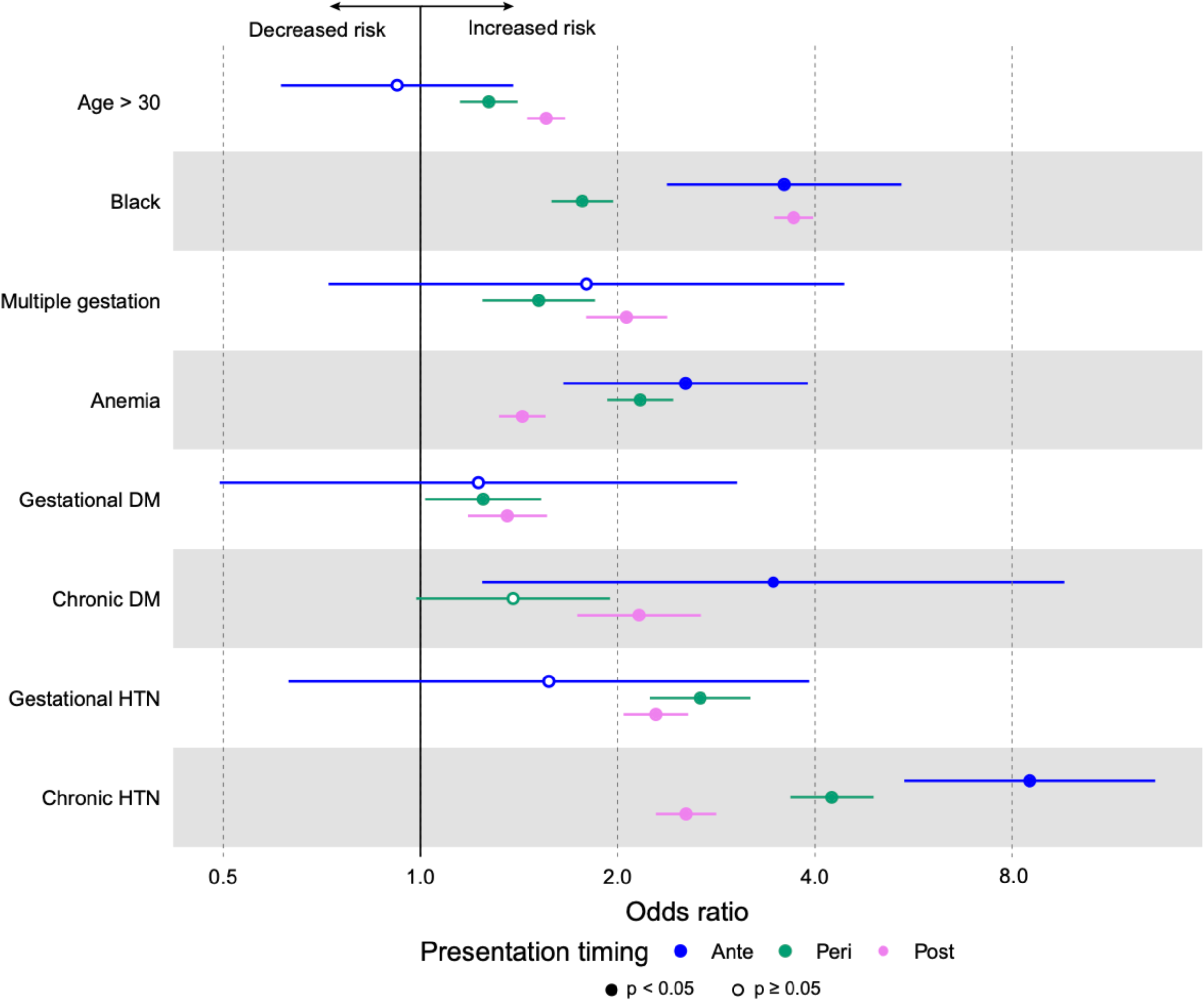
Multivariable odds of PPCM associated with maternal comorbidities. Forest plot displaying adjusted odds ratios (OR) with 95% confidence intervals for the association between baseline maternal comorbidities and PPCM, stratified by timing of presentation (antepartum, peripartum, and postpartum). Comorbidities examined include maternal age >30 years, AA race, multiple gestation, anemia, diabetes mellitus (gestational and chronic), and hypertensive disorders (gestational hypertension, chronic hypertension, and preeclampsia/eclampsia). Chronic hypertension and AA race were among the strongest predictors across all presentation timings. *Adjusted for age >30 years, race, multiple gestation, diabetes mellitus, and hypertensive disorders.

**Figure 3b.**
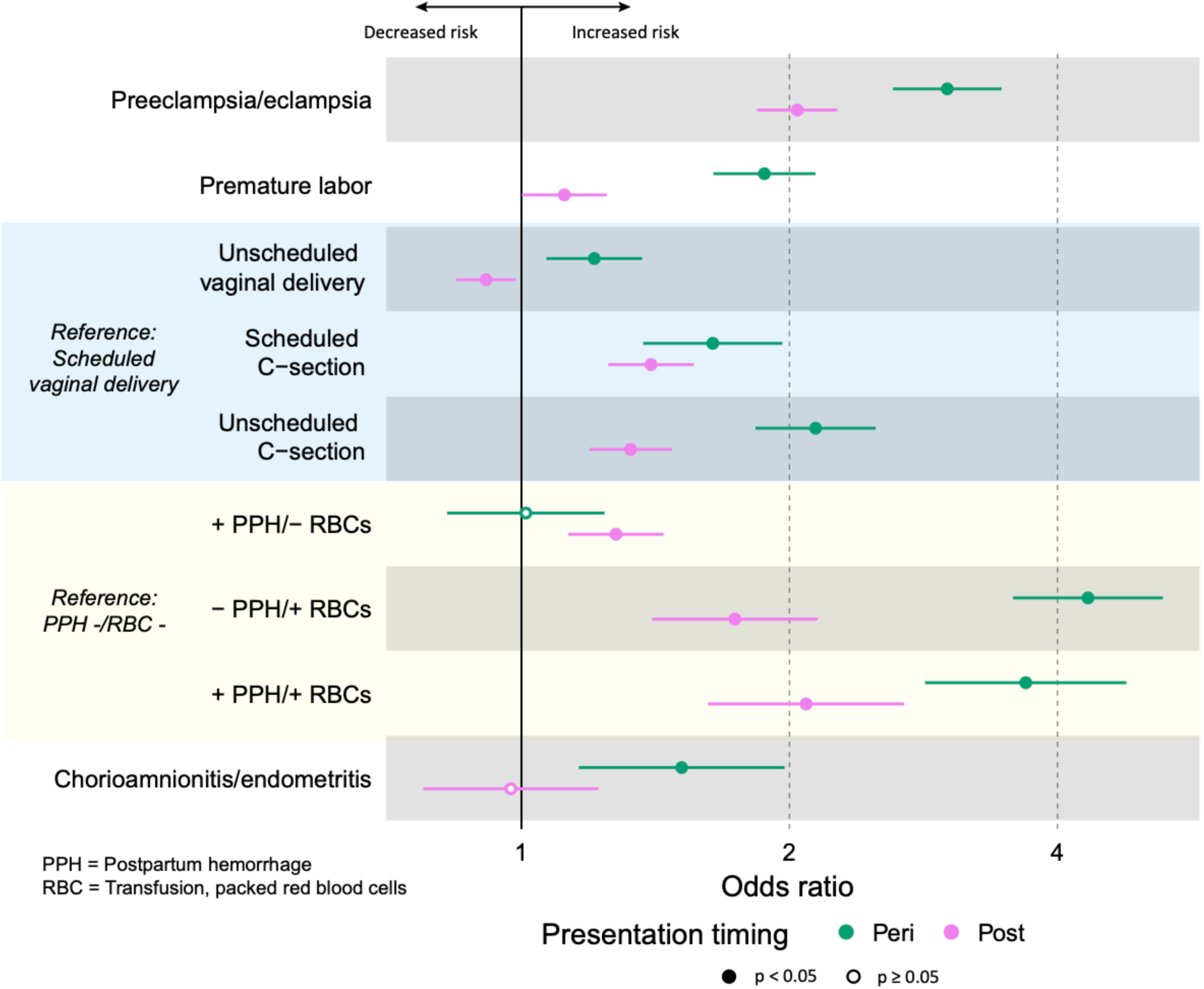
Multivariable odds of PPCM according to delivery events and complications. Forest plot showing adjusted odds ratios (OR) with 95% confidence intervals for the association between delivery-related events and complications and PPCM, stratified by peripartum and postpartum presentation. Delivery characteristics examined include premature labor, unscheduled vaginal delivery, scheduled and unscheduled cesarean section, postpartum hemorrhage, red blood cell (RBC) transfusion, and chorioamnionitis/endometritis. RBC transfusion without postpartum hemorrhage and unscheduled cesarean section were among the strongest predictors of peripartum PPCM. Reference categories were scheduled vaginal delivery and absence of postpartum hemorrhage or transfusion. *Adjusted for age >30 years, race, multiple gestation, diabetes mellitus, and hypertensive disorders.

Preeclampsia, premature labor, unscheduled delivery, blood transfusion, and chorioamnionitis were all associated with higher odds of peripartum PPCM compared with postpartum PPCM, whereas odds of postpartum PPCM was only nominally higher than peripartum presentation in post-partum hemorrhage only. Scheduled vs. unscheduled delivery was associated with increased incremental risk primarily in peripartum patients. PPH without RBC transfusion was associated with a slight increased risk for developing PPCM postpartum (OR 1.27, 95% CI 1.1-1.47). However, RBC transfusion both with and without PPH was associated with a markedly increased risk of peripartum PPCM (PPH+ OR: 3.68 [2.84-4.78] and PPH-: OR 4.33 [3.57-5.25], p < 0.001 for both) and postpartum PPCM (PPH+ OR: 2.09 95% CI: 1.62-2.69, PPH- OR: 1.74 [1.40-2.15], p < 0.001 for both). Chorioamnionitis/endometritis was associated with increased odds of peripartum presenting PPCM (OR 1.51 [1.16-1.98], p=0.002) but not postpartum presentation (p=0.813)

### Hemoglobinopathies and other anemia subtypes

On observing that RBC transfusion was associated with PPCM even in the absence of hemorrhage at delivery, we performed additional sub-analyses of anemia subtypes represented in the dataset. These included post-hemorrhagic anemia, anemia of chronic disease, iron deficiency anemia, sickle cell trait (SCT), sickle cell disease (SCD) and thalassemia. Other nutritional deficiencies, hemolytic, sideroblastic, and aplastic anemia were considered but there were too few events to analyze. We analyzed the full cohort as well as white and AA subsets due to the variable frequency of some hemoglobinopathies, most notably SCT and SCD. Although rates of coded anemia subtypes were generally low, in patients who developed PPCM, we observed significantly higher rates of post-hemorrhagic anemia (7.0% vs. 2.6%), anemia of chronic disease (ACD, 0.3 vs. <0.1%), IDA (3.5% vs. 1.2%), SCD (0.5% vs. 0.1%), SCT (1.6% vs. 0.6%), thalassemia (0.3% vs. 0.2%, p=0.013) and unspecified anemia (13.3%, 6.8%, p < 0.001 for all subtypes) (**Table 4**). Due to non-specific ICD coding, we were unable stratify by thalassemia phenotype. Comparatively low event rates limited comparisons based on PPCM timing, though in general associations seemed strongest in patients who developed PPCM antepartum. Trends generally held when stratified by white and AA race with most anemia subtypes being overrepresented in antepartum presenting patients. As expected, AA Patients with PPCM had higher rates of SCT and SCD than white patients. Among AA patients, both SCT and SCD were significantly more common in for all antepartum, peripartum, and postpartum presenting PPCM (SCT: 10.4% vs. 4.8% vs. 3.1%, p=0.011 and SCD: 4.2% vs. 1.5% vs. 0.7% respectively, p<0.001). There was a nominal association between SCT and peripartum PPCM in white patients, but this represented only 2 PPCM cases. Thalassemia rates did not differ between non-PPCM and Patients with PPCM in either white or AA patients though with very few cases (p=0.899 and p=0.317 respectively). Multivariable odds ratios for each anemia subtype adjusted for all risk factors described above including race are summarized in **Figure 4**. The strongest associations were between antepartum presentation and anemia of chronic disease (OR 16.3 [2.26-117.9], p=0.006), peripartum presentation and anemia of chronic disease (OR 10.3 [5.47-19.2], p<0.001) antepartum presentation and sickle cell disease (OR 9.11 [2.21-37.5], p=0.002). Again, multivariable odds ratios associated with anemia subtypes were highest in antepartum-presenting patients with PPCM and lowest or non-significant in postpartum presentations. **[Table 4]**

**Figure 4.**
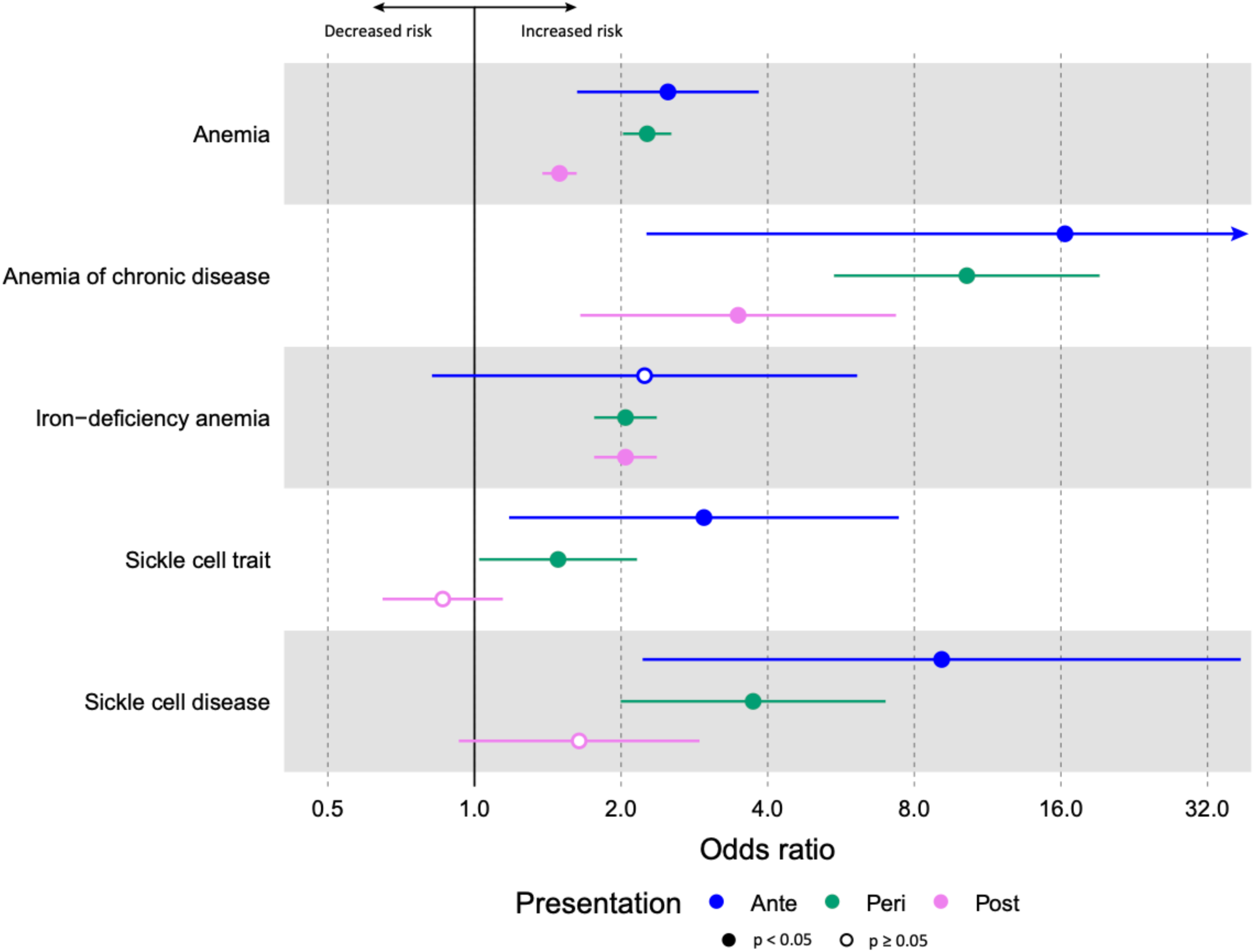
Multivariable odds of PPCM associated with anemia subtypes. Forest plot illustrating adjusted odds ratios (OR) with 95% confidence intervals for the association between specific anemia subtypes and PPCM, stratified by timing of presentation (antepartum, peripartum, and postpartum). Anemia subtypes evaluated include post-hemorrhagic anemia, anemia of chronic disease, iron-deficiency anemia, sickle cell disease, sickle cell trait, thalassemia, and unspecified anemia. Anemia of chronic disease and sickle cell disease demonstrated the strongest associations with PPCM across all presentation timings. *Adjusted for age >30 years, race, multiple gestation, diabetes mellitus, and hypertensive disorders.

**Table 4.**
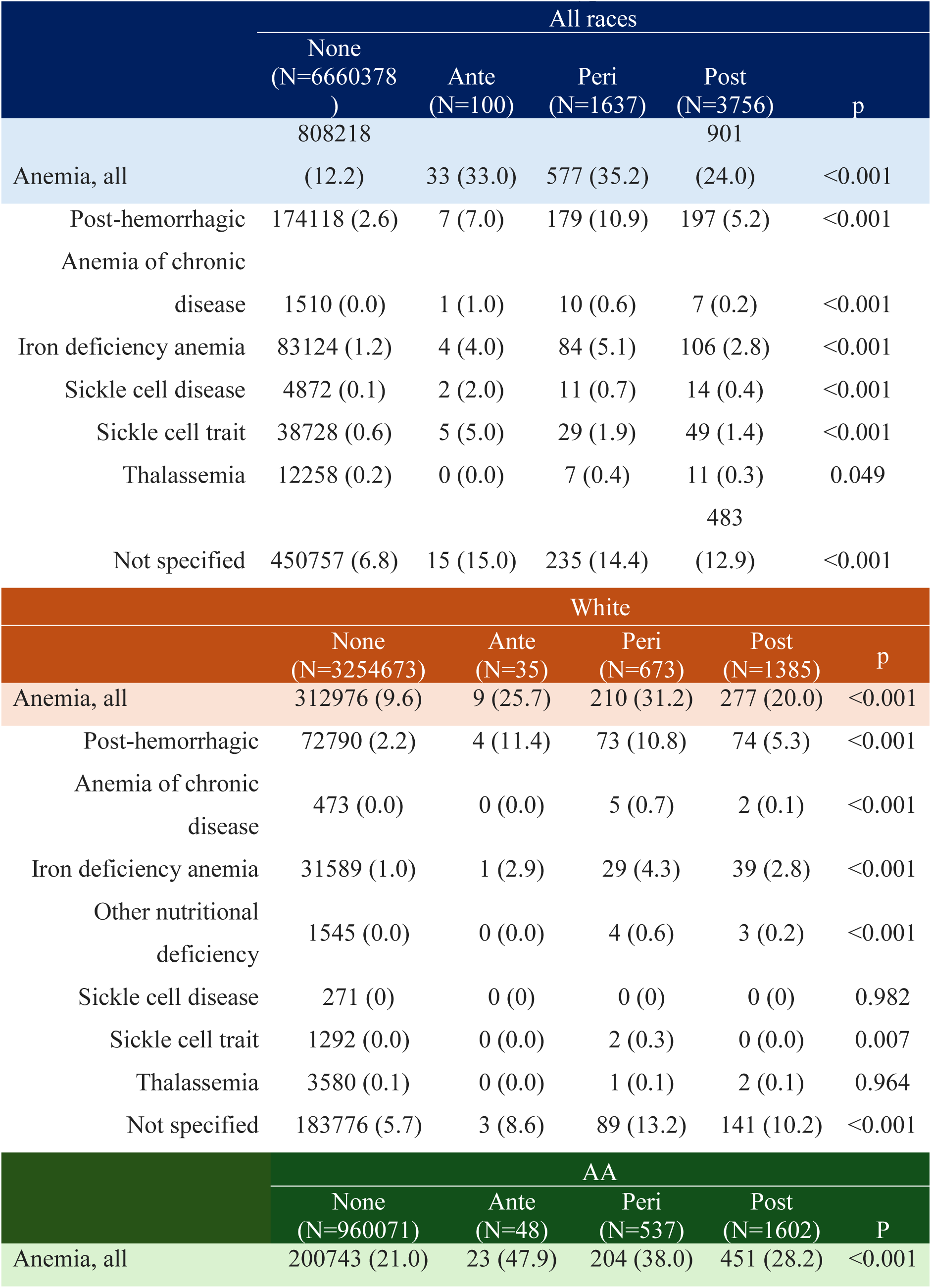
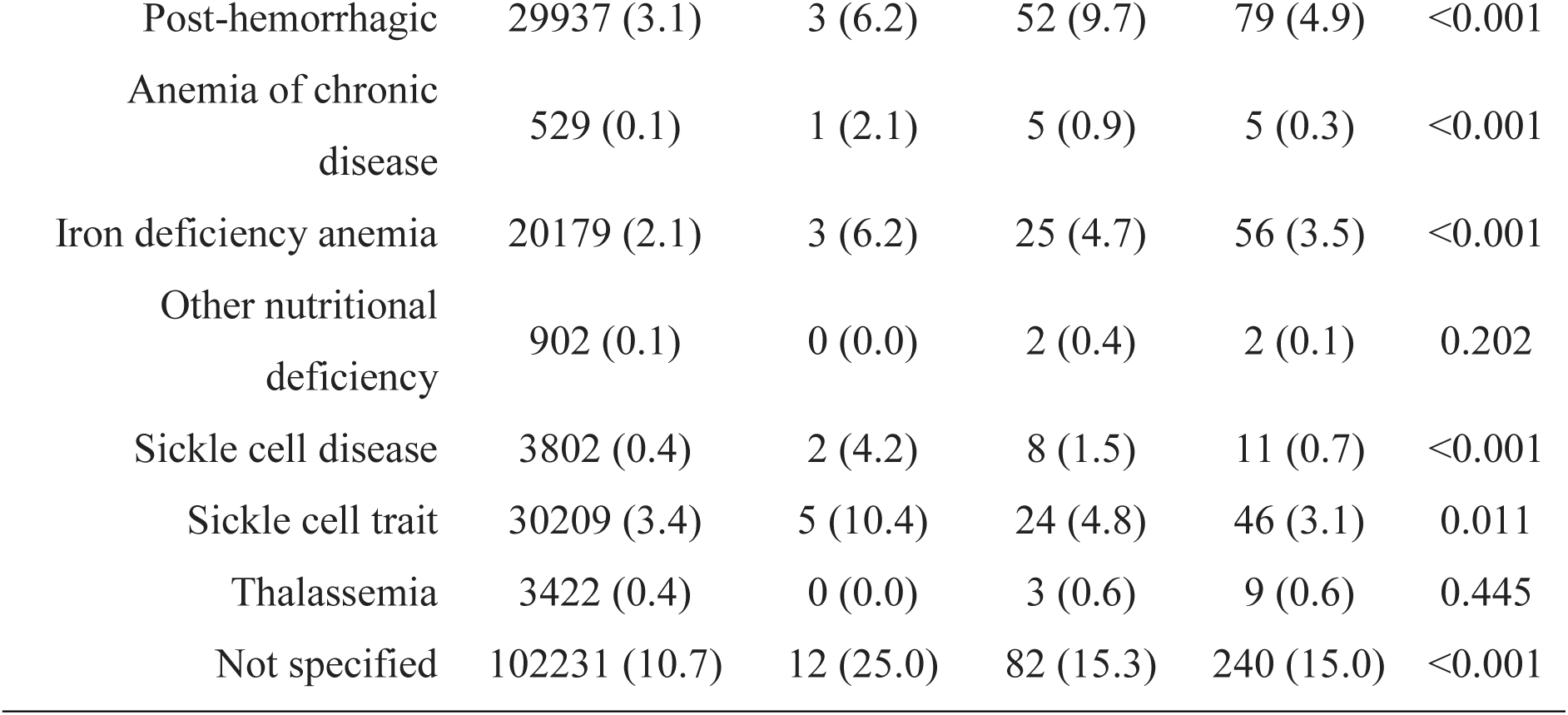
Association between PPCM and anemia subtypes.

## Discussion

Using a population of more than 6.6 million patients who have delivered with long-term readmission data, we found that complications at and immediately following delivery increase risk of subsequently developing PPCM independently of previously identified risk factors. We found that chronic forms of anemia and hemoglobinopathies including SCT were also associated with PPCM and transfusion of PRBCs at delivery was associated with a particularly high likelihood of presenting with PPCM in the postpartum period. We also found that patients with PPCM presenting antepartum had high rates of previously described risk factors including DM, cHTN, asthma, and SUD as well as higher rates of obstetric complications compared with those without PPCM, indicating that PPCM obstetric complications may be both a result of prior diagnosis and a risk for subsequent diagnosis of PPCM. Our findings suggest that obstetric course should be considered when assessing a high-risk patient’s risk of postpartum presentation with PPCM and a high index of suspicion should be used when evaluating nonspecific symptoms that could indicate heart failure such as lower extremity edema, dyspnea, and exercise intolerance.

While PPCM morbidity and mortality has improved over time due to reactive interventions including the ESC EORP PPCM recovery score to tailor heart failure treatment with bromocriptine and heart failure medications(32, 33), limited understanding of PPCM etiology has remained a barrier to identifying high risk patients for PPCM prevention and early intervention. Previous studies highlight the positive impact early diagnosis and intervention have on PPCM outcomes(34). However, the overlap between PPCM symptoms and physiologic symptoms of pregnancy and early postpartum, confounded by the lack of evidence-based clinical parameters and stratification tools to guide screening of high-risk patients, has made early diagnosis challenging. Timely workup is dependent largely on clinical suspicion supplemented in high-resource environments by B-type (BNP) and N-terminal pro-B-type natriuretic peptides (NT-BNP) blood tests, 12-lead ECG, and cardiac imaging such as transthoracic echocardiogram (TTE), followed by serial troponins at time of diagnosis(3, 4, 35). Access to cardiac imaging and biomarker testing can be limited in low resource settings(36), leaving clinicians to rely on risk assessment using historical data and physical examination. PPCM risk factors are heterogeneous, and multiple factors must be considered simultaneously(10, 33, 37). A clinical risk prediction model using historical data has been developed and validated (19) to prompt earlier diagnosis and intervention with a 3-5x increase in risk with each additional point with 12.5% of patients in the highest risk group (OR 37.55 and 25.17 compared with lowest risk patients in training and validation cohorts)(19). However, our prior prediction model has yet to be implemented in clinical practice, in part because optimal strategies for monitoring high-risk patients are unclear. We believe our observations on the importance of obstetric complications build upon our prior work by adding additional predictive information to existing models using data that is frequently available in the absence of sophisticated medical testing. As model performance improves in identifying extremely high risk patients, this will drive early detection, bridge racial and socioeconomic disparities, and improve morbidity and mortality for PPCM patients.

### Postpartum hemorrhage, anemia, and transfusion

PPH can be a severe complication of childbirth, with adverse outcomes ranging from prolonged hospitalization to need for blood products to surgical infertility via hysterectomy and even death. In this study, we found that PPH was associated with increased risk of PPCM, but we found that in multivariable analysis, the need for RBC transfusion seemed to drive the risk associated with PPH. Given that RBC transfusion also increased PPCM risk in the absence of PPH and that chronic anemia is one of the most significant risk factors for PPCM, our findings raise the question of whether a volume, vascular or oxygen-carrying capacity mechanism may be present and whether correction of anemia could be beneficial.

The American College of Obstetricians and Gynecologists (ACOG) defines IDA in pregnancy as a hemoglobin of less than 11g/dL and a ferritin less than 30ng/L (38). In 2023, IDA was established as a major public health concern by the World Health Organization (WHO), affecting 37% of pregnant patients worldwide(39) with higher prevalence in low and middle income countries (40). Importantly, severe IDA can lead to LV dysfunction and myocardial damage, contributing to heart failure development(41, 42), however the risk profile of iron deficiency in PPCM has not been investigated.

IDA has been associated with adverse pregnancy outcomes including preterm birth, PPH, increased maternal mortality, and cardiac complications (43). However, in 2024 the USPSTF reported that there was insufficient evidence to suggest there was clinical benefit to oral iron supplementation throughout pregnancy(44). Despite this, current CDC recommendations include daily supplementation of 27mg oral iron for all pregnant patients starting in the first trimester. Currently, there are multiple ongoing clinical trials investigating the safety and efficacy of IV iron repletion in iron deficient pregnant patients. Historically, IV iron use in pregnancy was limited due to high rates of adverse reactions with older formulations such as iron dextran, but these risks have been negated with modern formulations such iron gluconate. The RAPID-IRON trial investigated the use of IV iron in pregnancy and reported preliminary data that shows IV iron repletion in the second trimester decreased transfusion requirements and IV iron repletion in the third trimester in comparison to oral ferric carboxymaltose, without increased rates of adverse events(45). Our study found IDA to be associated with an increased risk for PPCM, with a stronger association observed in antepartum PPCM (OR 2.93 95% CI: 2.35-3.65) compared to postpartum PPCM (OR 1.64, 95% CI: 1.35-2.0) (p < 0.001 for both), irrespective of race (Table 3b). While the etiology of this association between IDA and PPCM is unclear, IDA may potentiate PPCM development through mismatch between myocardial oxygen delivery and demand, although it may be related to other social determinants of health such as optimal antepartum surveillance or ability to extend hospital stay related to anemia(8) (46). Ultimately, both IDA and RBC transfusion were associated with an increased risk for PPCM in this cohort. These findings, in conjunction with the RAPID-IRON trial findings, highlight the importance of future studies to characterize the cardiac implications of IDA in pregnancy and assess the efficacy of routine ferritin testing and IV iron repletion for patients with IDA to improve cardiac related maternal morbidity and mortality.

### Antepartum Screening

We noted that inherited forms of anemia were also associated with increased incidence of PPCM. Hemoglobinopathies and hemolytic anemias affect approximately 2.1 billion individuals globally, with an increased prevalence related to medical advancements and global migration(47). Current recommendations outlined by ACOG for anemia screening include a complete blood count, and RBC indices at the first antenatal visit(48). However, in August 2022, ACOG updated their recommendations to also perform hemoglobin electrophoresis in the first trimester to screen for genetic blood disorders regardless of ethnicity(48, 49). While various anemias and hemoglobinopathies have been associated with worse outcomes in heart failure with reduced ejection fraction (HFrEF) including iron deficiency(50–53), sickle cell disease(23), and thalassemia (54), these diseases have yet to be characterized in the context of PPCM. SCD has been associated with multiple complications of pregnancy that have been identified as risk factors for PPCM including African descent, anemia, preterm birth, and HDP (55). While SCD rates were higher in our PPCM cohort compared to patients without PPCM (0.5% vs. 0.1%, p< 0.001) (Table 3a), both SCD (OR 4.12, 95% CI: 2.32-7.3 p < 0.001), and SCT (OR 1.59, 95% CI: 1.13-2.25, p=0.008) were significantly associated with antepartum PPCM presentation only (Figure 5, supplemental table 2) when controlled for race. Meanwhile, SCD (OR 1.64 95% CI 0.93 - 2.91, p = 0.087) and SCT (OR 0.85 95% CI 0.65-1.15, p= 0.310) were not associated with an increased risk for developing postpartum PPCM (Figure 5, supplemental table 2). This data is limited by underdiagnosed SCD and SCT, and non-specific ICD anemia coding, hindering our ability to understand the true clinical significance and etiology behind this association. While pathways of vascular inflammation such as high output heart failure in the setting of chronic anemia, or cardiac remodeling due to chronic vaso-occlusion and inflammation should be considered, social determinants may also confound our findings. SCD and SCT primarily affect AA patients, who historically have more barriers to prenatal care(56), however patients with SCD and SCT are specifically recommended to be followed by both a general obstetrician and maternal-fetal medicine provider, significantly increasing their antenatal surveillance and potentially contributing to earlier detection of heart failure symptoms, though more studies are needed to characterize the true etiology of this association.

Thalassemia, to a lesser extent, is also associated with an increased risk for HDP and maternal anemia(57); however, this risk varies by phenotype and thalassemia rates did not vary between our non-PPCM and PPCM cohort, though these findings were limited by low power (Table 3a). Reassessment with larger cohorts following the update in ACOG recommendations for routine antenatal hemoglobin electrophoresis could provide a better understanding of the relationship between thalassemia, phenotype, race, and PPCM.

### Non-specific Implications of Peripartum Volume shifts

Physiologic changes of pregnancy, including fatigue, shortness of breath, and lower extremity edema overlap with acute heart failure symptoms, contributing to high rates of missed or delayed PPCM diagnosis(4). Throughout pregnancy, renin-aldosterone-angiotensin system (RAAS) activation leads to sodium retention and extravascular fluid shifts, and hypervolemia in the setting of RAAS dysregulation is a common cause of heart failure exacerbations(58–60) Intravascular fluid shifts and brisk diuresis take place within the first two weeks postpartum as atrial natriuretic peptide (ANP) increases, and estrogen, progesterone, aldosterone, and oxytocin decrease after delivery(61). Inhibition of ANP signaling in lactating mice has been associated with PPCM-like cardiac remodeling(62), and bromocriptine, a dopaminergic agonist with vasodilatory effects(63)has shown some promise in improving PPCM outcomes(5, 32) by inhibiting the cardiotoxic 16kDa prolactin fragment. Our study found that 84% of all Patients with PPCM presented within 20 days of delivery, within the window of postpartum fluid shifts (Figure 2). Additionally, RBC transfusion was found to be associated with an increased risk of PPCM regardless of hemorrhage status (Figure 3, supplemental table 1) suggesting the increased risk related to RBC transfusion may be related to volume status. These findings highlight the potential importance of vasodilation for diuresis and cardiac function in the immediate postpartum period. Despite these known physiologic changes of pregnancy that lead to increased cardiac demand, the effects of aggressive IV volume repletion in the postpartum period and the relation to pathogenesis of PPCM are unknown. Understanding the relationship between peripartum fluid shifts, IV volume repletion practices, and PPCM development and severity is critical for PPCM management and prevention.

## Limitations

Limitations of ICD-9 and ICD-10 code data analysis include coding inconsistencies, human error, and limited scope of exploration. Codes lack clinical specificity and granularity, often grouping diverse disease severities and subtypes under single codes while omitting crucial information about disease progression, severity, and treatment response. Specifically, differentiation between PPCM and hypertensive heart failure (HHFP) was more accurately defined in 2015, which may have led to inaccuracies in ICD coding and overrepresentation in this study(64). Financial incentives can systematically bias coding practices, with providers potentially over coding reimbursable diagnoses while underreporting clinically important but non-billable conditions. The coding system underwent periodic revisions and eventually transitioned to ICD-10, creating discontinuities that complicate longitudinal trend analysis. Furthermore, ICD data only captures what clinicians document and code, meaning undocumented or unevaluated conditions remain invisible in the dataset, and the absence of a code cannot reliably indicate the absence of a condition. Importantly, this data serves as a prompt for discussion and further investigation with either retrospective or prospective studies as granularity in care provided, such as total IVF administered, anti-hypertensive medication used, iron supplementation, or estimated blood loss cannot be determined with ICD codes and are essential in understanding the underlying pathophysiology of PPCM.

## Future Directions

Given the lack of clinical parameters to prompt diagnostic evaluation for PPCM, our lab has sought to validate a clinical risk prediction model. Previously identified risk clinical factors for PPCM include AA ancestry, genetic predisposition, age ≥ 30 years, HDP, cHTN, SUD, autoimmune disease, multiple gestations, asthma, mood disorders, obesity, diabetes mellitus, gestational diabetes, lower socioeconomic status and anemia(4, 6, 8, 10, 18, 19, 21). Previously, patients with 6 out of 9 of these characteristics had an 800-fold increase for developing PPCM(10). Davis *et al.* further developed and validated this model with a PPCM cohort of 1168 patients, finding that patients with 3 PPCM risk factors had a 28-fold increased risk of developing PPCM, while patients with 6 risk factors had a 189 fold increased risk of developing PPCM. These findings were further validated with our cohort and the incorporation of ICD-10 CM codes, suggesting patients with 3 or more non-genetic risk factors were associated with an 10.7 fold increased risk of developing PPCM (Table 1a). Recently, overall social vulnerability index was associated with worse morbidity and mortality in PPCM(22), therefore we aim to further investigate the association between PPCM, SVI themes and individual social risk factors with respect to race. This data will be used to to update and revalidate our proposed clinical risk stratification score with the novel risk factors identified in this study including RBC transfusion, PPH, IDA, SCD, unscheduled CD, ACD, and social determinants of health. The increased availability of EHRs, cloud platforms, and internet connectivity in the developing world also make possible broad dissemination of an updated risk model to the point-of-care, even in otherwise resource-limited settings.

The association between blood product transfusion with antepartum PPCM presentation, along with a majority of PPCM cases presenting within the window of postpartum fluid shifts highlights an understudied etiology that requires a diverse investigative approach. Better understanding of this association can guide evidence-based care, including correction of iron deficiency when present, judicious management of volume status through diuretics and minimizing transfusion of PRBCs. This may also support the utilization of ß-blockers such as carvedilol, in place of labetalol or nifedipine, the anti-hypertensives routinely used in pregnancy, given its superior vasodilatory effects, decreasing myocardial oxygen demand and cardiac strain as postpartum fluid shifts take place. highlights potential avenues for PPCM prevention in high-risk patients including, and judicious management of volume status through diuretics and minimizing transfusion of PRBCs. We next aim to explore the possibility of multiple mechanisms of PPCM development through deep phenotyping and association between true antepartum and postpartum presentation during delivery admission to determine when and in what context different mechanisms of PPCM are contributing.

## Conclusion

We identified novel risk factors that induce vascular stress including PPH, IDA, SCD, unscheduled CD, and RBC transfusion significantly increase the risk for PPCM. Additional studies are required to understand these associations, particularly in regard to vascular inflammation and volume status. These novel risk factors should be considered when assessing clinical suspicion and diagnostic evaluation for PPCM in a high risk patient. In symptomatic patients with elevated risk of PPCM, vasodilatory beta-blockers, IV iron replacement, modest volume resuscitation, and minimization of PRBC transfusion may all have a role in reducing risk of PPCM but future research would be needed to confirm these hypotheses.

## Data Availability

Analysis code and scripts are available on GitHub platform. Remaining ICD-9 and ICD-10 CM codes used for each diagnosis or procedure for the present analysis are available in the on-line supplement. Any additional data or information is available upon request.

## Acknowledgements

We extend our gratitude to the Ludeman Family Center for Women’s Health Research and the Jacqueline Marie Schauble Leaffer Endowment that made this research possible.

## Data availability statement

Analysis code and scripts are available on **GitHub** platform. Remaining ICD-9 and ICD-10 CM codes used for each diagnosis or procedure for the present analysis are available in the on-line supplement. Any additional data or information is available upon request.

